# Hospital wastewater surveillance for SARS-CoV-2 identifies intra-hospital dynamics of viral transmission and evolution

**DOI:** 10.1101/2025.03.04.25323323

**Authors:** Medini K. Annavajhala, Anne L. Kelley, Lingsheng Wen, Maya Tagliavia, Sofia Moscovitz, Heekuk Park, Simian Huang, Jason Zucker, Anne-Catrin Uhlemann

## Abstract

Wastewater testing has emerged as an effective, widely used tool for population-level SARS-CoV-2 surveillance. Such efforts have primarily been implemented at the wastewater treatment plants (WWTPs), providing data for large resident populations but hindering the ability to implement targeted interventions or follow-ups. Conversely, building-level wastewater data exhibits increased variability due to rapid daily population dynamics but allows for targeted follow-up interventions or mitigation efforts. Here, we implemented a three-site wastewater sampling strategy on our university-affiliated medical campus from May 2021 to March 2024, comprised of two distinct hospital quadrants and a primarily research laboratory and classroom building. We first addressed several limitations in implementing hospital-level wastewater surveillance by optimizing sampling frequency and laboratory techniques. We subsequently improved our ability to model SARS-CoV-2 case counts using wastewater data by performing sensitivity analyses on viral shedding assumptions and testing the utility of internal normalization factors for population size. Our unique infrastructure allowed us to detect intra-hospital dynamics of SARS-CoV-2 prevalence and diversity and confirmed that direct sequencing of wastewater was able to capture corresponding clinical viral diversity. In contrast, research building wastewater sampling showed that for most non-residential settings, despite low overall viral loads, a threshold approach can still be used to identify peaks in cases or transmission amongst the general population. Our study expands on current wastewater surveillance practices by examining the utility of and best practices for upstream and particularly hospital settings, enabling the use of non-municipal, medium-scale wastewater testing to inform efforts for reducing the burden of COVID-19.

**Importance:** Since the onset of the COVID-19 pandemic, wastewater surveillance has been increasingly implemented to track the spread of SARS-CoV-2. Most wastewater testing across the United States occurs at municipal wastewater treatment plants. Yet, this testing method could also be beneficial at non-municipal and non-residential sites, including hospitals, where wastewater data for SARS-CoV-2 signals and viral diversity could directly impact hospital practices to control its spread. We analyzed both hospital and non-residential research building wastewater over a three-year period to establish optimized methods for collecting and interpreting wastewater data at sites upstream of treatment plants. We found that even within a single hospital building, wastewater testing in different locations showed distinct signatures over time, which corresponded with data from patients hospitalized in those locations. This study provides a framework for the use of wastewater viral surveillance upstream of municipal treatment plants to enable targeted interventions to limit the spread of SARS-CoV-2.

## Introduction

Wastewater-based surveillance has been used since the onset of the COVID-19 pandemic to track and predict trends in SARS-CoV-2 cases at the community level.^1^ In the early pandemic phases, viral testing at municipal wastewater treatment plants (WWTPs) detected increases in estimated case counts earlier than clinical testing data,^2^ enabling public health messaging and awareness campaigns for regional case hotspots. Similar surveillance data from campus dormitories and other primarily residential settings has been used to detect individual cases and implement infection control practices such as quarantining or follow-up contact tracing.^3–5^ Wastewater testing is now considered the main tool available to estimate and predict the burden of SARS-CoV-2 in a given population due to highly variable SARS-CoV-2 testing practices and data availability.

As rates of clinical testing for SARS-CoV-2 decrease, hospital wastewater monitoring could provide aggregate data on estimated cases in not only patient but also visitor and staff populations. However, the implementation of this approach in hospital centers, unlike WWTPs and campus dormitories, has remained rare due to several unique challenges. First, wastewater data at the building level exhibits increased variability due to rapid daily changes in population size and dynamics, unlike larger scale monitoring at the WWTP level. Second, while hospital-level wastewater surveillance is still cost-effective compared to testing of individuals, the patient population at medical centers can be highly diverse, comprising inpatient units of varying care levels, outpatient clinics, and emergency departments. Specific viral variants have also been shown to impact on not only patient outcomes, but also factors which could impact wastewater testing, such as length of infection and viral shedding rates.^6^ However, most wastewater SARS-CoV-2 surveillance efforts focus on viral quantification rather than identification of viral lineages. Given these factors, data modeling and interpretation techniques used for large scale WWTP testing are not directly applicable to the unique aspects of smaller, upstream sampling scales.

Several strategies can be implemented to overcome challenges specific to hospital or non-residential wastewater testing. For example, increased sampling frequency and the use of internal population markers can account for rapid population fluctuations to improve accuracy of modeled case counts.^7–9^ Targeted or whole-genome sequencing of SARS-CoV-2 from wastewater can also be used to identify variant-specific dynamics and compare lineage prevalence within a hospital, for instance, versus the broader community.^10,11^ However, the impact of each of these approaches on the accuracy of SARS-CoV-2 prevalence or diversity estimates using wastewater testing data have not been systematically assessed, particularly in hospital or non-residential settings. Due to the availability of robust clinical testing and sequencing data,^12^ our hospital center represents a unique opportunity to study the ability of wastewater viral quantification and sequencing to accurately reconstruct clinical case counts and viral diversity.

In this study, we implemented wastewater sampling on our university-affiliated medical campus from May 2021 to March 2024. Wastewater testing was conducted at three sites, including two distinct hospital quadrants and a building with primarily research laboratories and classrooms, to determine intra-hospital dynamics of SARS-CoV-2, which differed in magnitude and diversity at various times throughout the sampling period. Comparison with clinical SARS-CoV-2 specimens linked by patient location was performed to determine if directly sequencing wastewater was sufficient to capture clinical viral diversity and evolution. We contrasted this approach with the utilization of wastewater data from our research building, reflecting testing realities in non-residential or commercial settings.

## Methods

### Wastewater surveillance infrastructure

This study was approved by the Columbia University Irving Medical Center Institutional Review Board (Protocol No. IRB-AAAU1682).

The primary adult inpatient hospital units at NewYork-Presbyterian Hospital Columbia University Irving Medical Center campus (NYP-CUIMC) are housed in a 10-story building organized in four vertical quadrants. Wastewater from floors 4-10 flows to one of four house traps located in the hospital ground floor, and then from each house trap into the city sewer system. Two of four hospital house traps were selected for ongoing surveillance, referred to in this study as Hospital Quadrant A and B (HQA, HQB). Sampling at HQA was initiated in May 2021, and at HQB in June 2021.

Our third selected sampling site, referred to as research building (RB), represents wastewater collected from above floor two of two connected buildings, both primarily comprised of research laboratories and classrooms. Sampling at RB was initiated in November 2021.

At each of the three sites, 24-hour composite samples were collected initially five times per week (May 2021 – August 2023) and subsequently three times per week (August 2023 – March 2024) except during necessary sampler maintenance. At sites HQB and RB, Teledyne ISCO Glacier refrigerated water samplers were installed for automated composite sampling; at site HQA, an in-house setup using an NCON Systems Sentry pump with attached refrigerator was used. Raw sewage was stored on-site at 4°C until transport to the laboratory for processing. An overview of all collected samples is available in **Supplementary Figure S1**.

### Viral concentration and nucleic acid extraction from wastewater

From May 2021 – January 2022, viral particles were concentrated from raw wastewater using polyethylene glycol (PEG) precipitation (see **Supplementary Methods**). PEG-precipitated viral pellets were resuspended in 0.3 mL of 1X phosphate buffered saline (PBS) and stored at -80°C. After January 2022, we used the InnovaPrep Concentrator Pipette for viral concentration (see **Supplementary Methods**). Two 50 mL aliquots of raw wastewater from each site were concentrated. One concentrated aliquot was stored without additional storage buffer, and the other was mixed with an equal volume of Zymo DNA/RNA shield for nucleic acid stabilization. Both aliquots were stored immediately at -80°C.

RNA was extracted using the RNeasy Mini kit (Qiagen), and both DNA and RNA were extracted using the Zymo DNA/RNA Miniprep Kit. DNA was immediately stored at –20°C. The LunaScript RT Supermix Kit (NEB) was used to generate cDNA from extracted RNA, stored at –20°C, and remaining RNA was stored at –80°C.

### Quantification of SARS-CoV-2 via quantitative reverse transcription polymerase chain reaction (qRT-PCR)

Quantification of SARS-CoV-2 was performed using qRT-PCR. Reactions were performed in duplicate using the 2019-nCoV RUO Kit (IDT) premixed primer and probe for N2 gene detection, following the CDC-recommended protocol^13^ (see **Supplementary Methods**). A standard curve was generated for each assay using the 2019-nCoV_N_Positive Control plasmid (IDT) at 50 ge/μL to 4,000 ge/μL. Each assay included at least two no-template negative controls.

### Quantification of CrAssphage via quantitative polymerase chain reaction (qPCR)

CrAssphage was quantified in duplicate via quantitative polymerase chain reaction (qPCR) on DNA extracted from each wastewater sample. Primer and probe sequences and reaction setup followed methods described in ^14^ using the referenced CPQ_056 primers and probe (see **Supplementary Methods**). Each assay included a standard curve at 10-fold dilutions spanning from 5 ge/μL to 500,000 ge/μL and at least two no-template negative controls.

### Calculation of SARS-CoV-2 and CrAssphage viral load

Viral concentrations in terms of copies per mL wastewater were calculated for both SARS-CoV-2 and CrAssphage as follows (**Equation 1**):

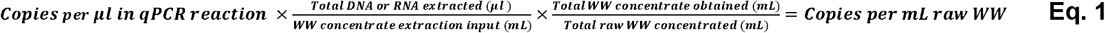

### Data imputation and cross-correlation

Time-lag cross-correlation was performed to test for significant association between wastewater qPCR results from hospital sampling sites HQA and HQB (in terms of both weekly median cycle threshold value (Ct) or weekly median viral concentration (copies / mL wastewater)) and confirmed patient cases in each relevant hospital quadrant (see **Supplementary Methods** for details on inpatient SARS-CoV-2 case count data and assumptions for patient viral shedding dynamics). Weekly sampling was near-complete through the study period; in HQA, 7 of 149 (5%) potential sampling weeks were missed, and in HQB, 10 of 144 (7%) potential sampling weeks were missed. Prior to cross-correlation, data imputation for these missing values was performed using local rolling median values (k=5, *zoo* R library). The cross-correlation function (*ccf*) in R was then used to compare cleaned wastewater and clinical case data and obtain R^2^ values to describe the strength of each time-lag cross-correlation.

### Whole-genome and targeted gene sequencing of SARS-CoV-2 from wastewater

Samples with an average Ct below 33 cycles were selected for whole-genome sequencing of SARS-CoV-2. Tiled amplification of the viral genome was performed using the xGen SARS-CoV-2 Amplicon Panel (IDT). Sequencing libraries were prepared from amplicons with the Oxford Nanopore Native Barcoding Expansion 96 kit (NBD110.96 or NBD114.96), and 15-50 fmol of pooled barcoded libraries were then sequenced on an Oxford Nanopore GridION sequencer using R9.4.1 or R10.4.1 flow cells.

Nanopore adaptive sampling was used to align reads to the SARS-CoV-2 genome (NCBI RefSeq NC_045512.2) in real-time during sequencing, to enrich for target library fragments. All reads were then re-mapped to the NC_045512.2 reference genome using *minimap2* with default parameters for long-read mapping. A BED file with genomic coordinates and primer sequences for the xGen SARS-CoV-2 Amplicon Panel was used to identify primer binding sites and filter mapped variants and generate masked BAM files. To identify the distribution of SARS-CoV-2 lineages in each sample, variants were called from BAM files using *ivar*^15^, and converted to variant frequency tables using *freyja*.^11^ A minimum coverage of 10X as calculated by *freyja* was required for inclusion of a sample in our analysis. Outputs from *freyja* were imported to R for visualization.

### Whole-genome sequencing of SARS-CoV-2 from clinical nasopharyngeal swabs

Nasopharyngeal swabs from patients testing positive for SARS-CoV-2 were sequenced as previously described (see ^12^ and **Supplementary Methods**). Briefly, cDNA from positive clinical specimens underwent tiled whole-genome amplification and library preparation using the Midnight Expansion Kit and Rapid Barcoding Kit (RBK110.96, Oxford Nanopore), and were sequenced on an Oxford Nanopore GridION with R9.4.1 flow cells. Genome alignment, consensus sequence generation, and lineage calling were performed via the Epi2Me wf-artic Nextflow pipeline (Oxford Nanopore).

## Results

### Hospital and research building surveillance overview

Over the nearly three-year study period between May 2021 to March 2024, a high frequency of dense wastewater sampling was achieved (**Supplementary Figure S1**).

**Supplementary Figure S1.**
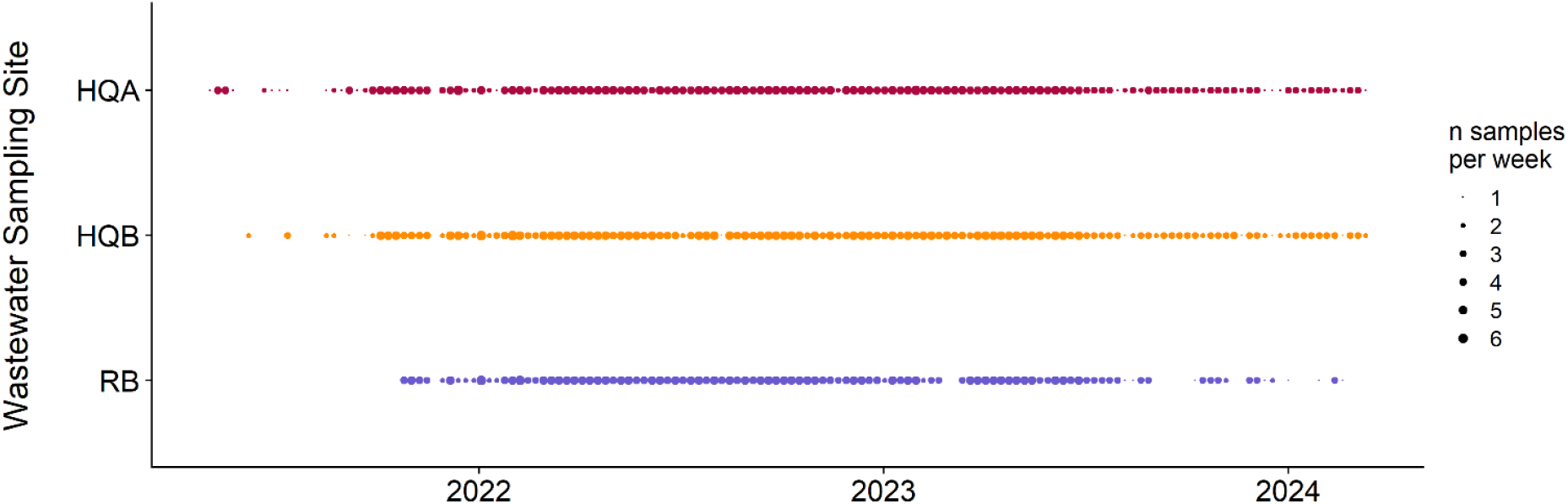
Hospital and research building sampling overview. For each sampling site (y-axis) over each week of our sampling period (x-axis) from May 2021 through March 2024, points are scaled by the number of samples collected per week. Abbreviations: HQA = hospital quadrant A, HQB = hospital quadrant B, RB = research building

We aimed to collect multiple samples per week from each site to test the impact of sampling frequency on the correlation between wastewater viral concentration and confirmed SARS-CoV-2 clinical cases. Sampling frequencies tested ranged from one to five times per week. In total, HQA was sampled for a total of 149 weeks, HQB for 144 weeks, and RB for 121 weeks (**Supplementary Table S1**). Missing data due to sampler outage and/or staffing changes was minimal in HQA and HQB, representing 7 (5%) weeks and 10 (7%) weeks, respectively. At least three samples per week were collected for HQA in 113 of 149 (76%) weeks, at HQB for 112 of 144 weeks (78%), and at least five samples per week were collected for HQA in 54 (38%) and for HQB in 55 (37%) weeks.

The sampler in our research building encountered significantly more outages or missed collections due to low, intermittent daily flow rates compared to the hospital samplers HQA and HQB. Despite this, samples were still collected for most of the study period: at least once weekly for 107 of 121 weeks (88%), at least thrice weekly for 90 weeks (74%), and at least five times weekly for 49 weeks (41%). Due to the lack of accurate control data in terms of confirmed SARS-CoV-2 cases, as staff were not routinely nor comprehensively tested throughout the study period, wastewater viral levels in the RB sampler could not be used for modeling or sensitivity analyses, but rather as an indicator of peaks in cases within the broader non-patient population as described in detail below.

**Supplementary Table S1.**
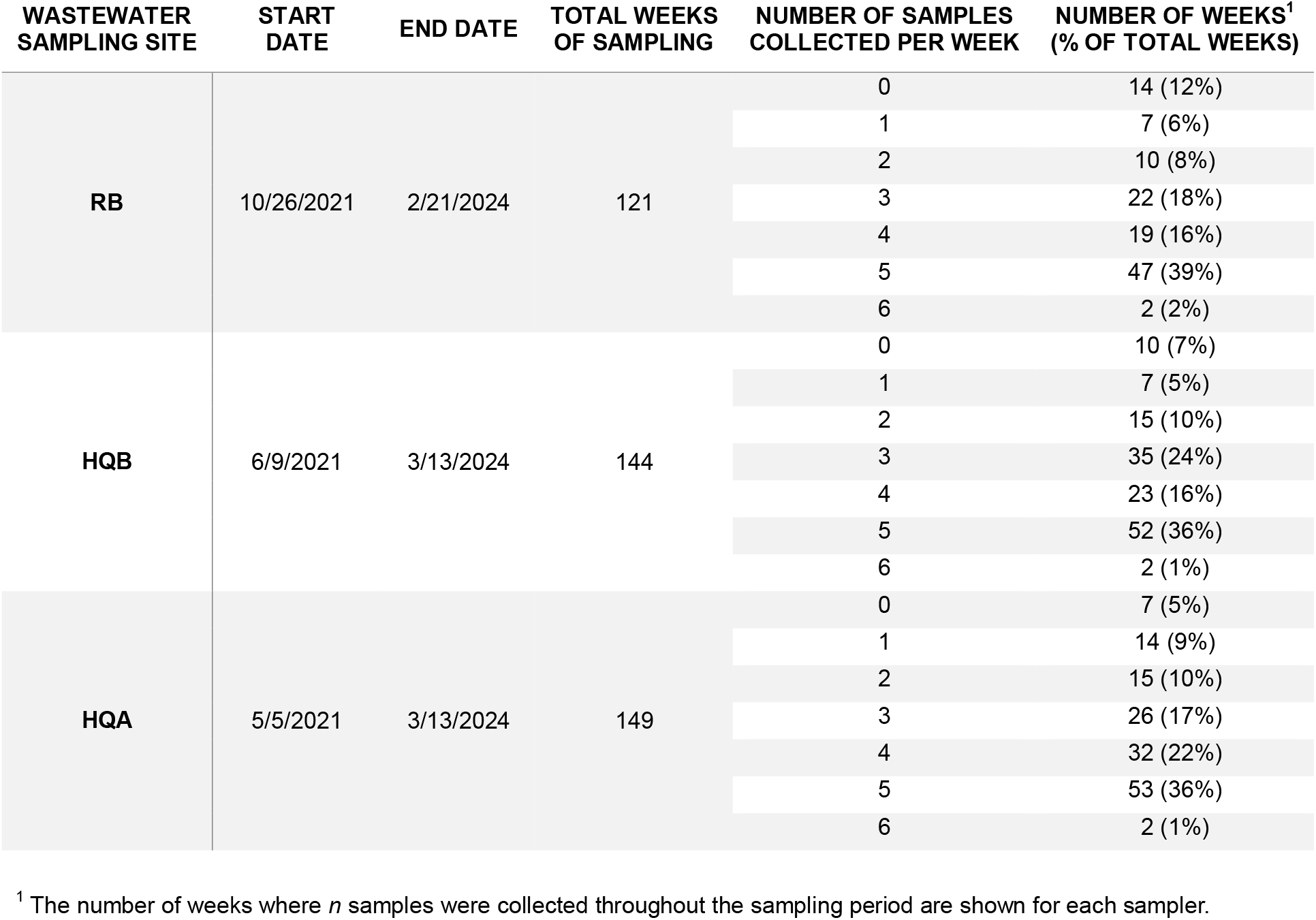
Weekly wastewater sampling frequency.

### Correlating wastewater SARS-CoV-2 concentrations with clinical case counts

The primary aim of this study was to establish the use of hospital or sub-hospital (quadrant) wastewater testing to monitor SARS-CoV-2 cases. This would allow individual hospital centers to move beyond the existing regional wastewater surveillance efforts at municipal wastewater treatment plants across the United States to enable targeted mitigation approaches only as needed.

We first established the verified SARS-CoV-2 clinical case data for each hospital quadrant sampled, HQA and HQB, longitudinally over our sampling period (**Figure 1**). Due to the sparse literature on gastrointestinal shedding rates and dynamics in individuals, we calculated the case counts assuming shedding windows of 3, 7, 10, or 14 days for each patient after their initial positive clinical test.

**Figure 1.**
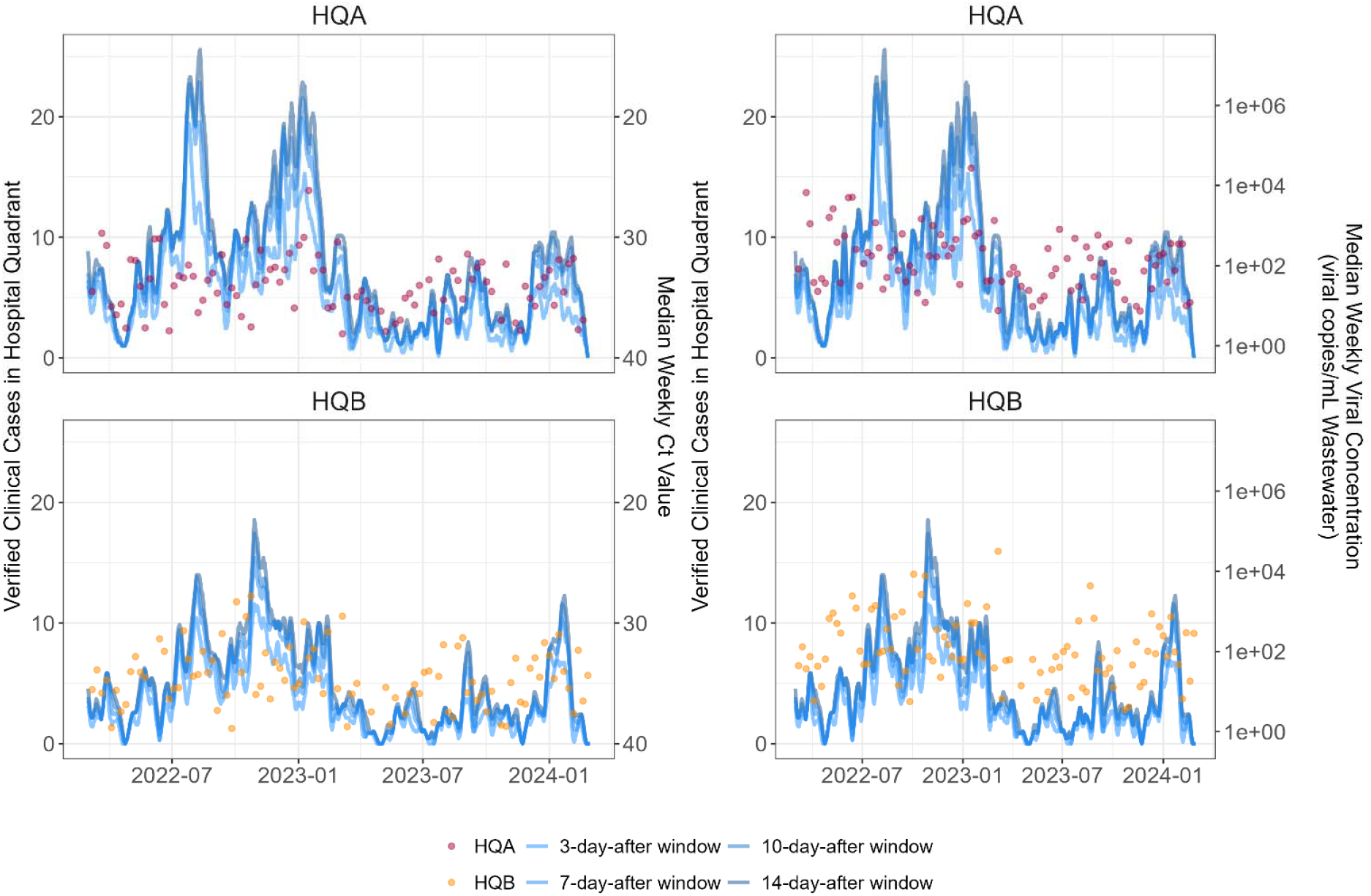
Wastewater viral concentrations and corresponding verified clinical cases throughout study period. Pink and orange dots show wastewater data from HQA and HQB, respectively. Blue lines show verified clinical patient cases of SARS-CoV-2 in the corresponding quadrants; from lightest to darkest, lines assume 3, 7, 10, or 14-day shedding windows for each patient, where day 0 is the date of the first positive clinical test. Panels on the left show the median weekly cycle threshold (Ct) values while panels on the right show median weekly viral concentrations (viral copies per mL raw wastewater).

We ran a series of lead-lag correlation analyses to test the strength of correlation between our upstream wastewater viral data and confirmed clinical cases, and to test whether our dataset exhibited a similar lead time as previous reports.^2^ Viral concentrations in copies per mL raw wastewater, both with and without including prior case count data, were compared with hospital quadrant cases assuming 3, 7, 10, or 14 days of gastrointestinal shedding for each patient after the first clinical positive test (**Table 1**; **Supplementary Table 2**). The rationale for the wastewater data only and wastewater and clinical data models was to mimic real-life scenarios where either only wastewater data, for hospitals without routine SARS-CoV-2 case tracking, or where both types of data would be available, as at our hospital center.

**Table 1.**
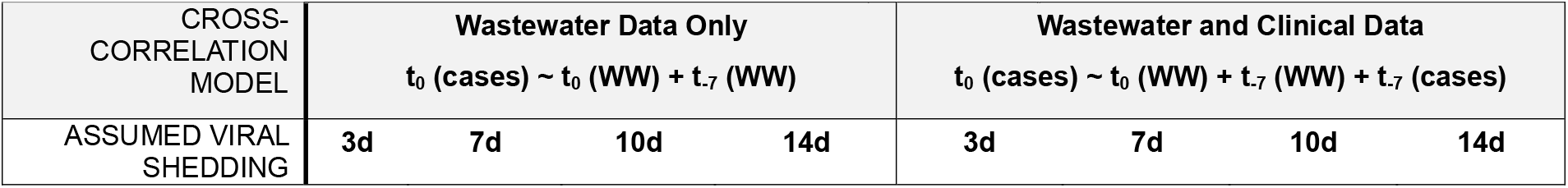
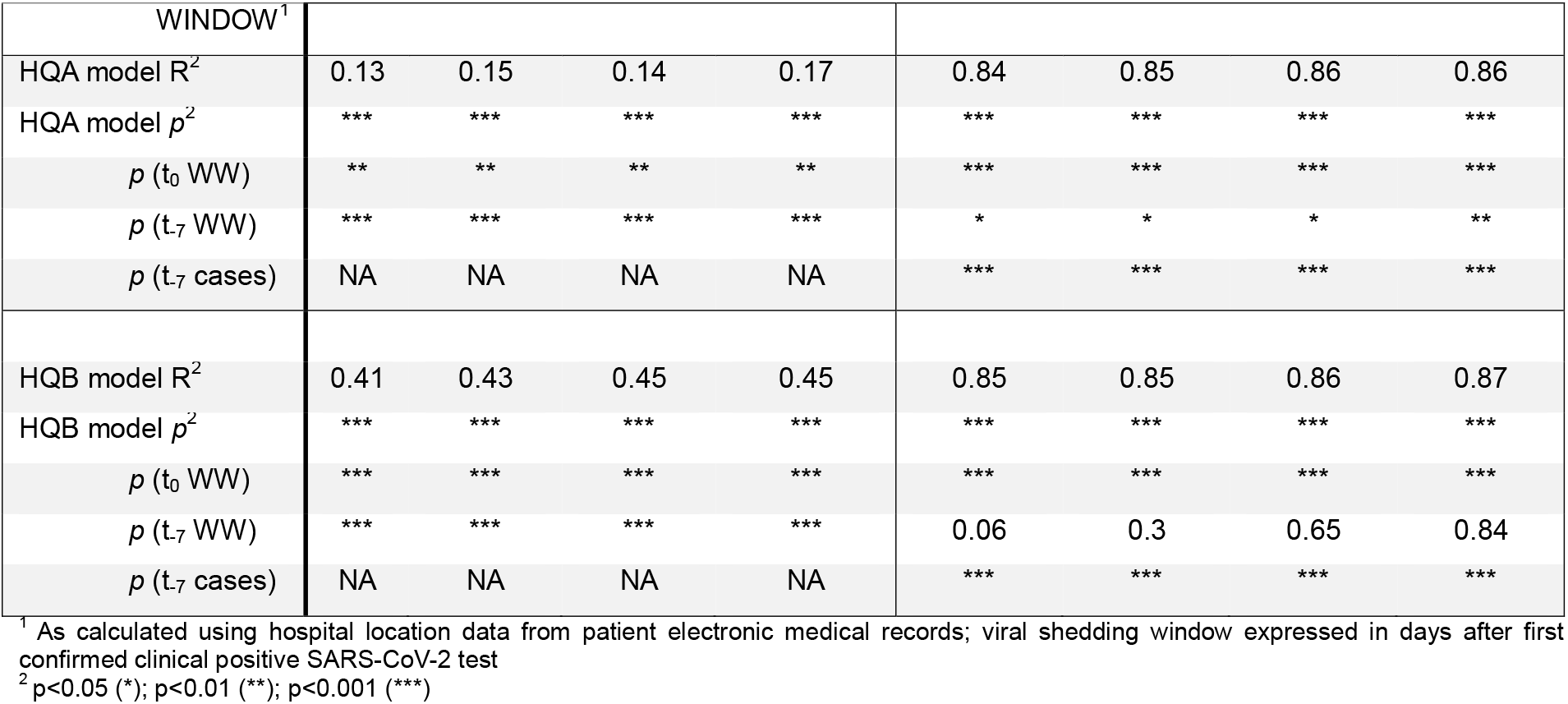
Lead-lag correlation analysis between wastewater viral concentration and confirmed clinical cases with shedding dynamics sensitivity analysis.

When using wastewater data only, we found the strongest statistically significant correlation between wastewater viral concentration observations at t_0_ and one week prior (t_-7_) to a given observation of cases (t_0_ (cases)) assuming a 14-day shedding window for HQA (R^2^ = 0.17) and to an even greater degree HQB (R^2^ = 0.45) (**Table 1**). The strength of the correlation increased further when including case observations from one week prior as a term in the multiple regression model (t_0_ (cases) ∼ t_0_ (wastewater viral concentration) + t_-7_ (wastewater viral concentration from 7 days prior) + t_-7_ (cases from 7 days prior); **Table 1**). We next used 60 weeks of consecutive data from HQA with concurrent SARS-CoV-2 and crAssphage viral concentration data available. Unexpectedly, normalization with crAssphage did not significantly impact the strength of correlation nor lead time for viral load data (R^2^ for 7-day lag time with 14 day assumed shedding= 0.32 with normalization versus 0.33 without normalization; **Supplementary Table S2**).

**Supplementary Table S2.**
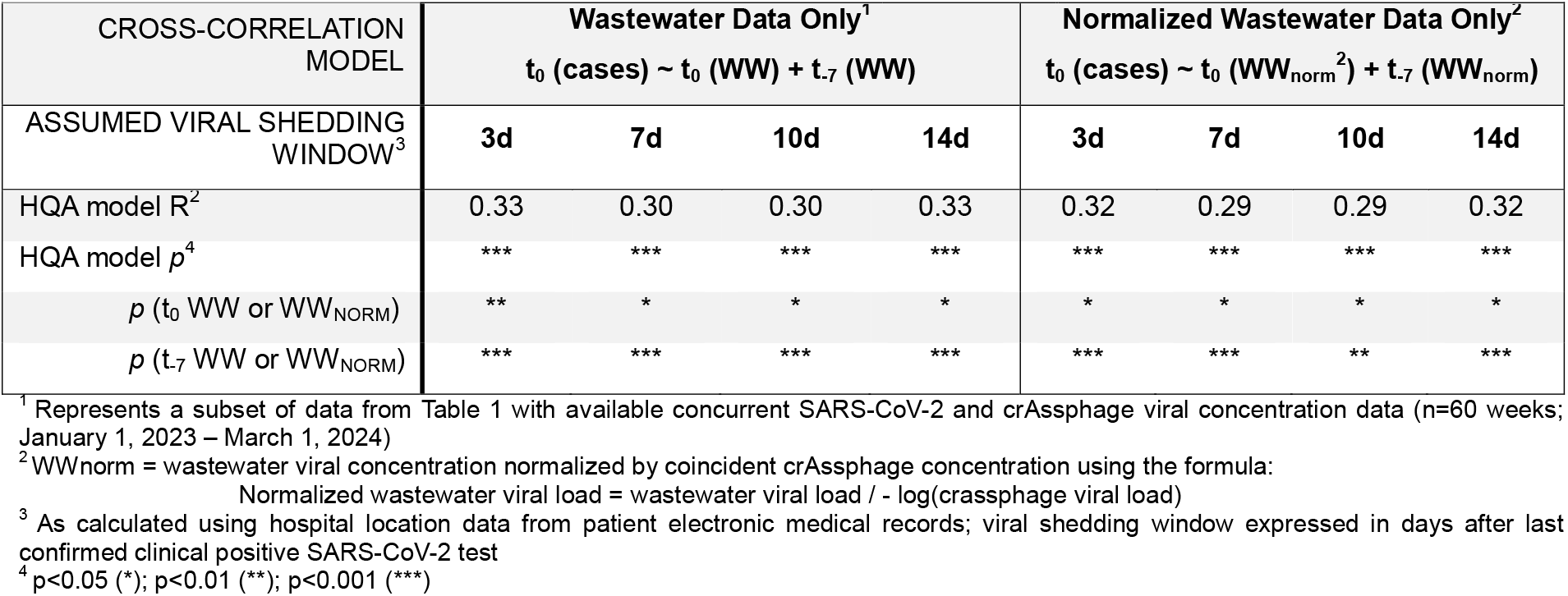
Lead-lag correlation analysis between wastewater viral concentration and confirmed clinical cases in HQA with shedding dynamics sensitivity analysis, with and without normalization by crAssphage concentration.

We also aimed to determine the minimum weekly sampling frequency required to maintain a significant correlation and lead time between wastewater viral data and confirmed cases. Data was first subset to between October 1, 2021 to March 1, 2023, when both sites were sampled at least 3 times for the majority of weeks (thrice-weekly sampling for 78% of weeks within timeframe for HQA, 60% for HQB) (see **Supplementary Figure S1** and **Supplementary Table S1**). For each week in this time period, selection of 1, 2, or 3 observations was randomized and used to re-calculate median weekly viral concentration, which was then correlated with clinical cases as above. We found that thrice-weekly sampling of HQA was needed at minimum to maintain a significant correlation with clinical cases, while weekly sampling was sufficient for HQB (**Supplementary Table S3**).

**Supplementary Table S3.**
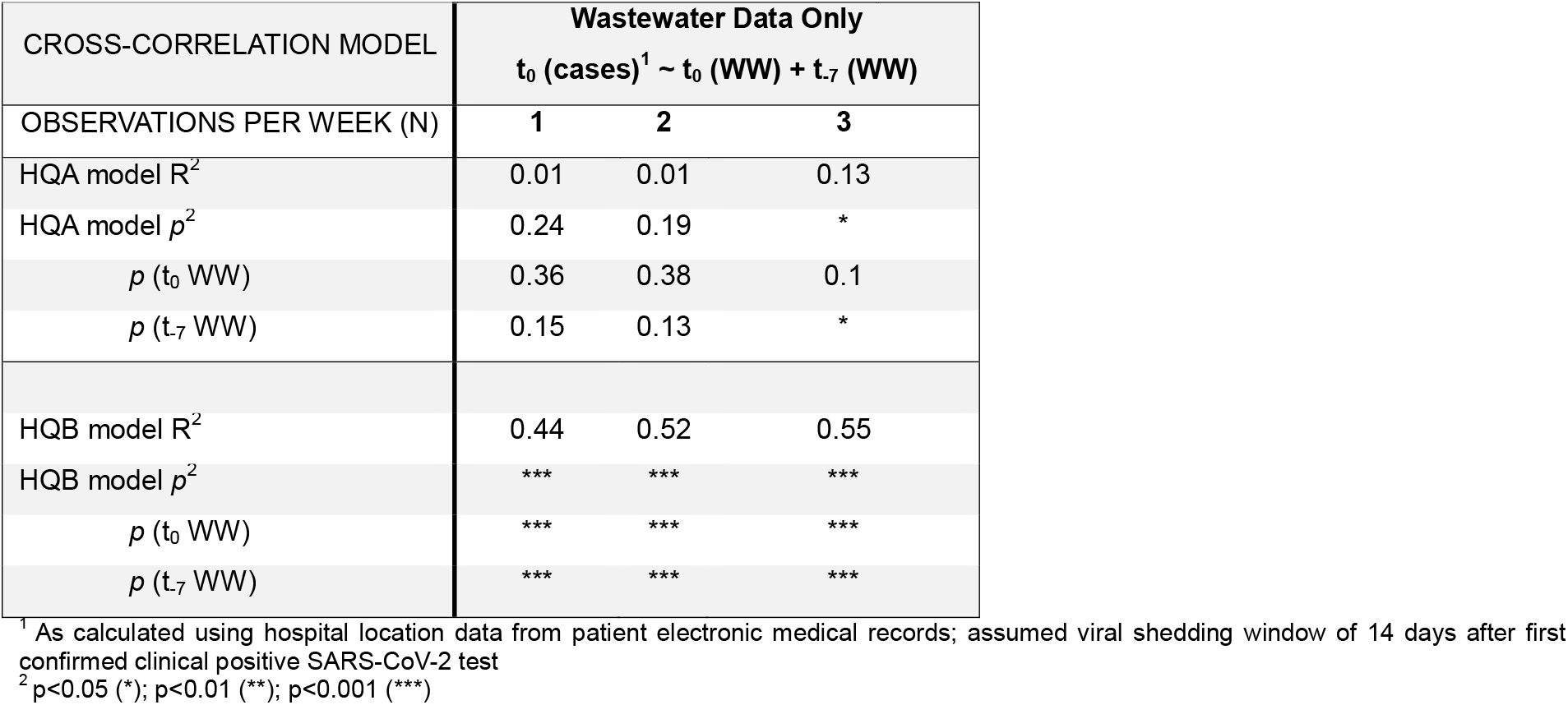
Lead-lag correlation analysis between wastewater viral concentration and confirmed clinical cases with sampling frequency sensitivity analysis.

| CROSS-CORRELATION MODEL | Wastewater Data Only |  |  |
| --- | --- | --- | --- |
| | $t_0$ (cases)<sup>1</sup> ~ $t_0$ (WW) + $t_{-7}$ (WW) | | |
| OBSERVATIONS PER WEEK (N) | 1 | 2 | 3 |
| HQA model $R^2$ | 0.01 | 0.01 | 0.13 |
| HQA model $p^2$ | 0.24 | 0.19 | * |
| $p$ ($t_0$ WW) | 0.36 | 0.38 | 0.1 |
| $p$ ($t_{-7}$ WW) | 0.15 | 0.13 | * |
| HQB model $R^2$ | 0.44 | 0.52 | 0.55 |
| HQB model $p^2$ | *** | *** | *** |
| $p$ ($t_0$ WW) | *** | *** | *** |
| $p$ ($t_{-7}$ WW) | *** | *** | *** |

<sup>1</sup> As calculated using hospital location data from patient electronic medical records; assumed viral shedding window of 14 days after first confirmed clinical positive SARS-CoV-2 test
<sup>2</sup> $p < 0.05$ (\*); $p < 0.01$ (\*\*); $p < 0.001$ (\*\*\*)

### Intra-hospital dynamics in wastewater SARS-CoV-2 diversity

We next aimed to investigate whether whole-genome sequencing of SARS-CoV-2 from wastewater would reflect the same variant profiles as our ongoing genomic surveillance of patients hospitalized in each corresponding hospital quadrant. Wastewater samples with Ct<33 were selected for whole-genome sequencing (n=133 samples from HQA and 99 from HQB). 104 of 133 (78%) samples from HQA and 73 of 99 (74%) samples from HQB met the criteria of 10X genomic coverage over at least 80% of the SARS-CoV-2 genome after sequencing. Relative frequencies of major variants as calculated using Freyja were plotted longitudinally for each hospital quadrant. Our ongoing genomic surveillance of all SARS-CoV-2-positive clinical nasopharyngeal swabs at our hospital center with Ct_≤_30 allowed us to directly compare wastewater variant frequencies not only across hospital quadrants but also with viral diversity from patients hospitalized in these quadrants (**Figure 2**).

**Figure 2:**
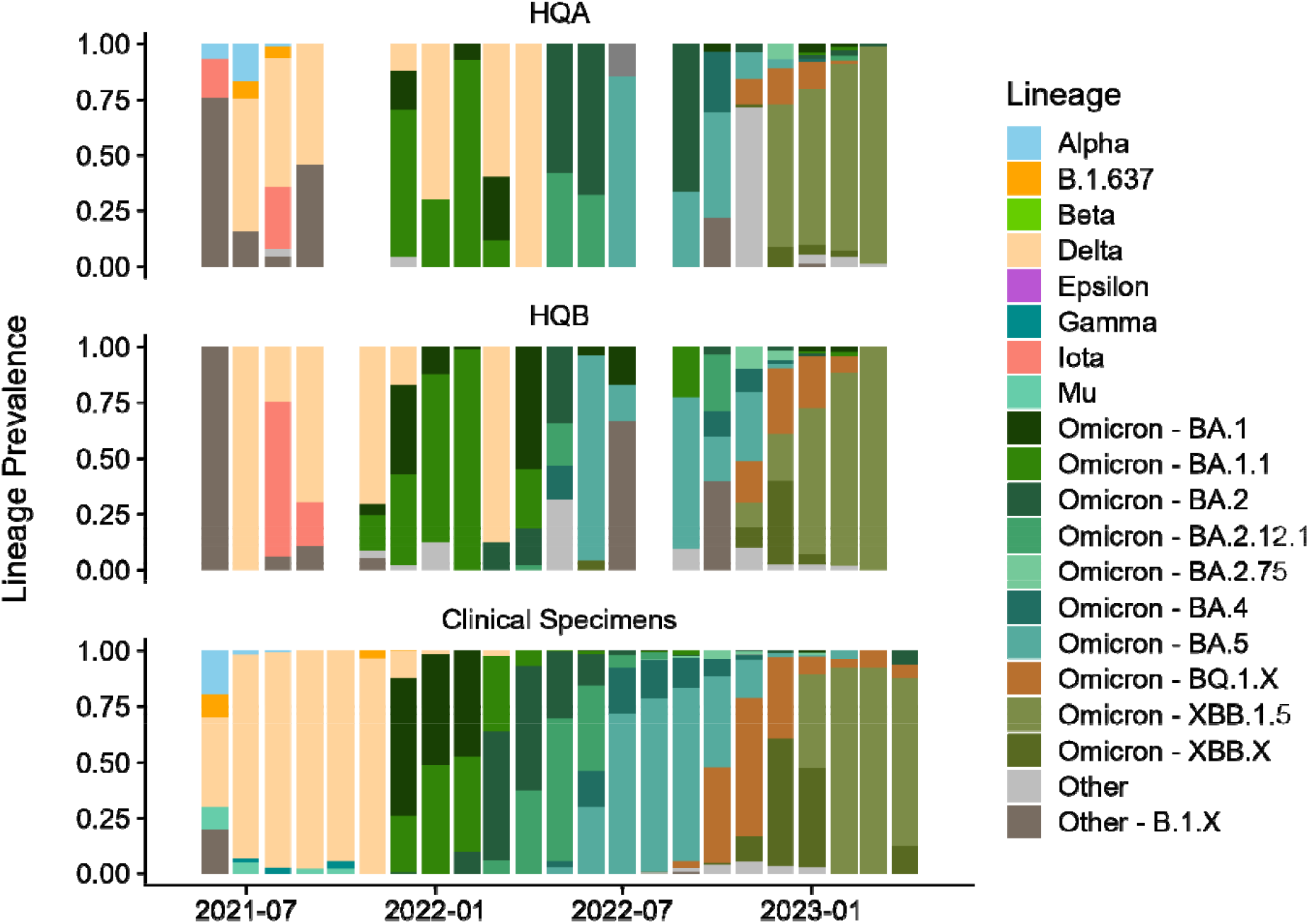
Whole-genome sequencing of SARS-CoV-2 from wastewater and clinical samples across hospital quadrants. Viral variant diversity in hospital quadrants HQA and HQB (**top, middle**) and from patients hospitalized in HQA or HQB during date ranges with contemporaneous wastewater sequencing data (**bottom**). Bar plots show variant frequencies along the sampling time period from samples with Ct<33. Wastewater viral lineage frequencies were largely concordant with clinical specimen sequencing data, with several notable exceptions where wastewater data revealed distinct spatial dynamics of specific SARS-CoV-2 lineages within the hospital environment (e.g., Alpha present in HQA but not HQB in mid-2021), indicated likely introduction or expansion of lineages not captured by clinical data (e.g., Delta in early 2022; recombinant lineages labeled “Other” in HQB in mid-2022), or provided evidence of lineage emergence prior to clinical sequencing efforts (e.g., XBB.1.5 in late 2022 to early 2023).

While longitudinal trends in both HQA and HQB wastewater sequencing reflected general trends observed in the patient population, distinctions between quadrants and between wastewater and clinical data were observed (**Figure 2**). For example, the Alpha variant was detected in HQA wastewater and in clinical specimens from June through August 2022, but not in HQB wastewater, pointing to the specificity of each quadrant’s wastewater sequencing data. Wastewater sequencing also revealed likely introductions or expansions of certain lineages that were not reflected in clinical specimen data. For example, the Delta variant was observed in both wastewater quadrants in early 2022 despite the dominance of Omicron BA.1 then BA.2 in the patient population during this timeframe. Similarly, the HQB quadrant wastewater contained evidence of recombinant lineages absent in both HQA and clinical specimens in mid-2022. Lastly, hospital wastewater from both quadrants showed the presence of emerging lineages in advance of clinical genomics efforts, as with XBB.1.5 observed in wastewater from both quadrants in late 2022 but not in clinical data until early 2023.

### Non-residential campus building surveillance reflects known SARS-CoV-2 case peaks, but with limited sensitivity

Sampling at the research building RB was performed as a case study to test the utility of wastewater-based surveillance in a non-residential, mixed-use setting. At the beginning of the sampling period, most staff had resumed in-person work after the initial pandemic shutdown. SARS-CoV-2 testing was performed primarily using random biweekly sampling of a subset of employees accessing RB through mid-2021. Thus, unlike for hospital or municipal wastewater testing, particularly during the initial two years of the pandemic, RB represents a setting where no comprehensive case data were available.

We compared wastewater viral concentrations in RB from Fall 2021 to Spring 2024 with longitudinal clinical case count data (**Figure 3**). For most weeks, the RB sampler showed no detectable viral load (SARS-CoV-2 qPCR Ct > 40 in 68 of 107 weeks with at least one sample collected, 64%). For the 39 of 107 weeks (36%) with detectable viral copies, concentrations were generally low, with median raw weekly Ct value of 36.4 (IQR 35.5-37.5) representing 66 (IQR 24-250) viral copies per mL raw wastewater.

**Figure 3.**
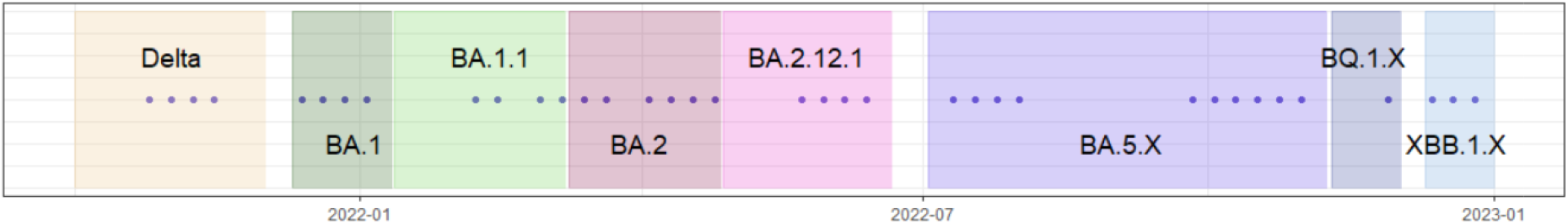
Weeks with detectable SARS-CoV-2 levels at RB site. Weeks with detectable viral concentration are shown as points through the sampling period for the research building RB. In total, 39 of 107 weeks with samples collected (36%) had a SARS-CoV-2 concentration greater than 0 copies / mL raw wastewater. Each cluster of detected positives coincided with a known surge in community cases based on publicly available data and data from our own hospital center, as labeled in the image. Shaded regions indicate date ranges where each variant or variant cluster was dominant (defined as comprising at least 50% of all sequenced cases as part of our routine genomic surveillance of all SARS-CoV-2 cases at our hospital center).^12^

Despite low viral concentrations in RB wastewater, weeks with a detectable signal corresponded with known case peaks based on hospital surveillance, obtained from our ongoing genomic surveillance efforts,^12^ and comparison with public health data from New York City, when available (**Figure 2, Figure 3**).

## Discussion

Our three-year wastewater surveillance effort at an urban university-affiliated hospital center was aimed at addressing existing gaps in implementing pathogen detection at sites upstream from municipal wastewater treatment plants. Through dense longitudinal sampling over the study period and extraction of patient location data from electronic medical records, we were able to establish cross-correlation models which allow for the prediction of SARS-CoV-2 case counts in each independent hospital quadrant using either wastewater data only or a combination of wastewater and clinical data, from up to one week before an observed increase in clinical cases. We also established that sampling at our hospital center scale did not benefit from normalization of wastewater viral concentration data using the internal population marker crAssphage, as has been previously described.^7,8,14^ Wastewater surveillance at the sub-hospital scale, providing quadrant-level data, allowed us to identify unique dynamics in viral levels and diversity between different hospital quadrants. Lastly, while wastewater sampling in our research building exhibited low overall viral loads, we were able to apply a threshold approach to identify peaks in cases or transmission amongst the building population.

Our data show that increases in hospital wastewater SARS-CoV-2 concentrations do significantly correlate with future increases in patient cases, preceding cases by up to one week, even at the hospital quadrant level. This relationship is observed when there is access to only wastewater data but is significantly strengthened when historic patient case data is also available, as is the case in hospital systems. These models also do not require normalization of wastewater data despite increased day-to-day variability in the patient, visitor, and staff population at a hospital building compared to a residential area, simplifying the laboratory workflow and analysis pipelines. However, NYP-CUIMC is a major urban hospital center; assessment of normalization factors for wastewater testing at medical facilities with significantly lower patient and staff census would be beneficial in future studies.

While sampling at the hospital quadrant level still represents a significant cost-, material-, and time-saving method compared to testing of every individual, we observed that at least thrice weekly sampling may be required to maintain statistically significant correlations between wastewater and case data, unlike the commonly implemented weekly surveillance at municipal treatment facilities. Uniquely, unlike municipal treatment facilities, surveillance at hospitals or individual buildings (e.g., employee buildings or universities) has the added advantage of a targeted population for which clearly identified decision-makers are empowered to quickly implement strategies to mitigate viral spread. Existing hospital pipelines for infection prevention and control could be leveraged in the same way using targeted wastewater data showing the current and upcoming case burdens within a single hospital center or even a single hospital section.

Both viral levels and SARS-CoV-2 variant diversity were distinct across the two hospital quadrants tested, pointing to the ability of increasingly targeted wastewater testing to isolate spatial trends within a single hospital center. Importantly, sequencing of the SARS-COV-2 genome directly from wastewater was able to recapitulate viral lineage frequencies observed via genomic surveillance of individual patient samples. Thus, while genomics of individual clinical samples may be indicated for the purposes of clinical care, general longitudinal surveillance of frequently occurring or novel SARS-CoV-2 variants could be achieved through wastewater sequencing. This approach would allow for decreased cost, labor, and turnaround times by sequencing aggregate quadrant-level wastewater instead of individual clinical samples. Our data also provides evidence for the ability of wastewater sequencing to identify emerging viral lineages prior to their detection in clinical samples. A limitation of this approach is that a relatively low cycle threshold value of Ct 33 is generally required for successful whole-genome viral sequencing; during periods of low overall case burden, alternative methods may be required to capture viral diversity.

Wastewater surveillance has not been routinely established in mixed-use non-residential buildings due to unique challenges, as substantiated in this study. Fluctuations in the building population throughout the week, or even during a single day as in the case of periodic classroom usage, can lead to low wastewater flow rate and highly variable wastewater testing data. The disposal of waste from laboratories within the building into a combined water and sewer system can also be expected to impact the detection of pathogens in wastewater. However, despite these challenges, we were able to detect SARS-CoV-2 during periods of known case peaks as determined via public health databases and our own hospital surveillance. Hence, while a quantitative model of case burden was not possible at this site, presence of SARS-CoV-2 signal in this mixed-use, non-residential building could be used to detect periods of significant case burden by applying a simple threshold reporting method (detected versus non-detectable) rather than requiring absolute quantification of SARS-CoV-2 virus.

While the data presented here strengthen the case for hospital wastewater monitoring of SARS-CoV-2 and establish various best practices for implementation, several limitations of this study must be noted. The hospital sites tested here represent a snapshot of our large tertiary care medical center. Specific targets including intensive care units or hospital units with large proportions of immunocompromised patients, which may exhibit unique viral case dynamics, was not possible given the existing infrastructure. Therefore, implementing hospital-wide wastewater surveillance systems may be constrained by engineering or architectural concerns, or the added cost these may entail. Methodology implemented in this study may or may not be applicable to efforts extending beyond SARS-CoV-2 to other clinically relevant pathogens; further studies will be needed to establish whether pathogen-specific protocols are required to establish more comprehensive hospital pathogen wastewater surveillance.

In sum, our study examined the utility of and best practices for wastewater monitoring upstream of regional wastewater treatment plants, particularly in hospital settings. These findings will help enable the use of non-municipal, medium-scale wastewater testing for more targeted and effective mitigation efforts to reduce the burden of COVID-19 and can be applied in the future to diverse pathogens of interest. Future work to bridge the pipeline from wastewater data to administrative mandates, established in conjunction with hospital or commercial decision-makers, would further facilitate effective use of wastewater surveillance data beyond municipal public health efforts.

## Data Availability

All sequencing data in FASTQ format is available via the NCBI Sequencing Read Archive (SRA) under BioProject PRJNA1230707.

## Funding

This work was supported by the National Institute on Drug Abuse (NIDA U01DA053949 to ACU and MKA) and the National Institute of Allergy and Infectious Diseases (NIAID K99AI163348 to MKA).

## Acknowledgments

We would like to acknowledge Dr. Brian Mailloux and the Barnard College, Columbia University, and NewYork-Presbyterian/Columbia Irving Medical Center Facilities teams for their advice and assistance in setting up wastewater surveillance infrastructure for this study. We would also like to acknowledge Dr. Eldad Hod and the Columbia University Biobank (CUB) team for providing clinical SARS-CoV-2 specimens for sequencing. CUB is supported by the Irving Institute for Clinical and Translational Research (NCATS UL1TR001873).

## Notes

### Competing Interest Statement

The authors have declared no competing interest.

### Author Declarations

The Institutional Review Board of Columbia University Irving Medical Center gave ethical approval for this work (Protocol No. IRB-AAAU1682)

