## Supplementary Methods for "Hospital wastewater surveillance for SARS-CoV-2 identifies intra-hospital dynamics of viral transmission and evolution"

+ Current affiliation and address:

Medini K. Annavajhala, Children’s Hospital of Philadelphia, Colket Translational Research Building, 3501 Civic Center Boulevard, Room 10010, Philadelphia, PA, 19104

\# Corresponding authors:

Anne-Catrin Uhlemann, MD, PhD,

Medini K. Annavajhala, PhD,

**Keywords**: wastewater surveillance, SARS-CoV-2, COVID-19, hospital wastewater, wastewater-based epidemiology

**Viral concentration and nucleic acid extraction from wastewater:**

*Polyethylene glycol (PEG) precipitation:* Two aliquots of composite raw wastewater were concentrated from each site. 6.25 mL of glycine buffer (0.05M glycine, 3% beef extract, pH 9.6) was added to 43.75 mL raw wastewater for a total volume of 50 mL and centrifuged at 5,000-8,000 x g for 30 min at 4°C. The supernatant was removed using a serological pipette to avoid disturbing the pellet and passed through a 0.2 or 0.45 µm polyethersulfone filter. The filtrate was then incubated overnight with agitation at 4°C with 5X 1M PEG 8000 and 1.5M sodium chloride. The next day, the sample was centrifuged at 13,000 x g for 2 hours at 4°C. The supernatant was discarded, and the pellet was resuspended with 0.3 mL of 1X phosphate buffered saline (PBS) and stored at -80°C.

*InnovaPrep Concentrator Pipette*: Two 50 mL aliquots of composite raw wastewater from each site were centrifuged at 4,500 x g for 15 min at 4°C to remove debris. The supernatant was decanted into a fresh 50 mL tube, and 500 µL of 10% Tween20 was added. Samples were then filtered through an Innoveprep Concentrating Pipette Select using the ultrafiltration pipette tip (~0.01 µm pores) and 25 mM Tris FluidPrep Elution Fluid. One concentrated aliquot was stored without additional storage buffer, and the other was mixed with an equal volume of Zymo DNA/RNA shield for nucleic acid stabilization. Both aliquots were stored immediately at -80°C.

*Nucleic acid extraction:* For early samples from May 2021 – February 2022, RNA was extracted using the RNeasy Mini kit (Qiagen) with an input of 150 µL concentrated wastewater in 1:1 DNA/RNA Shield, and an elution volume of 60 µL. From January 2022 – March 2024, the Zymo DNA/RNA Miniprep Kit was used to isolate both DNA and RNA from 175 µL of wastewater concentrate in 1:1 DNA/RNA Shield and eluted in 75 µL RNA or 60 µL DNA. DNA was immediately stored at –20°C. The LunaScript RT Supermix Kit (NEB) was used to generate 30 µL single-stranded cDNA from 24 µL extracted RNA, which was stored at –20°C. Remaining RNA was aliquotted and stored at –80°C.

**Quantification of SARS-CoV-2 via qRT-PCR:** qRT-PCR reactions were performed in duplicate using the 2019-nCoV RUO Kit (IDT) premixed primer and probe for N2 gene detection, following the CDC-recommended protocol^1^: 5 µL of RNA or cDNA was added to 15 µL of reaction mix composed of 5 µL of 4X TaqMan Fast Virus 1-Step Master Mix (Thermo Fisher), 2 µL N2 primer probe mix, and 8 µL H_2_O. Amplification was performed as follows: 50°C for 5 min, 95°C for 20 s, and 40 cycles of 95°C for 3 s followed by 60°C for 30 s. A standard curve was generated for each qRT-PCR assay using the 2019-nCoV_N_Positive Control plasmid (IDT) at concentrations spanning from 50 ge/µL to 4,000 ge/µL. Each assay included at least two no-template negative controls.

**Quantification of CrAssphage via qPCR:** CrAssphage was quantified in duplicate via quantitative polymerase chain reaction (qPCR) on DNA extracted from each wastewater sample. Primer and probe sequences and reaction setup followed methods described in ^2^. 2 µL of DNA was added to 23 µL of reaction mix composed of 2X TaqMan Environmental Master Mix (Fisher Scientific), 1 µM forward and reverse CPQ_056 primers, 10 pM CPQ_056 probe, 5 µg bovine albumin serum (BSA, Thermo Scientific), and 1 µL H_2_O. The qPCR amplification was performed as follows: 95°C for 10 min followed by 40 cycles of 95°C for 15 s and 60°C for 1 min. A standard curve was generated using an in-house puc19 plasmid containing the crAssphage_056 insert as described in ^2^, at 10-fold dilutions spanning from 5 ge/µL to 500,000 ge/µL.

**Hospital system and inpatient SARS-CoV-2 case data:** Throughout the duration of this study, hospitalized patients at CUIMC/NYP were routinely tested for SARS-CoV-2 upon admission or clinical suspicion, or prior to surgical or other hospital procedures. The admission-discharge-transfer (ADT) dataset was extracted from the electronic health records of patients hospitalized at NYP-CUIMC from May 1, 2021 to present. Based on this dataset, hospitalization time of a patient was defined as starting from emergency department triage time to discharge time from the hospital (if applicable), and patients with at least one hour of hospitalization time were included in the analysis. In addition, a patient’s location inside the hospital every day was defined as their location at 8 pm of that day during their hospitalization, which was used to subset all hospitalized patients to those hospitalized in quadrants HQA or HQB.

**Viral shedding dynamics:** SARS-CoV-2 virus and/or viral RNA may be shed into wastewater starting from several days prior to confirmation of COVID-19 infection, and lasting days after a confirmed positive clinical test.^3,4^ We applied a time window of virus shedding for each COVID-19 positive case hospitalized in units located within quadrant HQA and HQB, from 2 days before to 3, 7, 10, or 14 days after the first confirmed positive clinical SARS-CoV-2 PCR test during the hospitalization. Using this data, we estimated the 7-day rolling average number of total patients, as well as COVID-19 cases, in each hospital quadrant over time.

**Whole-genome sequencing of SARS-CoV-2 from clinical nasopharyngeal swabs:** Nasopharyngeal swabs from patients testing positive for SARS-CoV-2 were sequenced as previously described.^5^ Briefly, after undergoing SARS-CoV-2 testing by the NYP-CUIMC Clinical Microbiology laboratory, positive specimens were inactivated using an equal volume of Zymo DNA/RNA Shield and stored at –80°C by the Columbia University Biobank. Samples were then requested by our laboratory and underwent RNA extraction using the QIAmp 96 Viral RNA kit on a QiaCube HT (Qiagen) using the CDC-recommended protocol,^1^ with an input of 70 µL sample in 1:1 shield and elution volume of 100 µL. The LunaScript RT Supermix kit was used to generate cDNA, which then underwent tiled whole-genome amplification and library preparation using the ONT Midnight Expansion Kit and Rapid Barcoding Kit (RBK110.96, Oxford Nanopore). Sequencing libraries were pooled and sequenced on an Oxford Nanopore GridION with R9.4.1 flow cells. Genome alignment, consensus sequence generation, and lineage calling via Pangolin were performed via the Epi2Me wf-artic Nextflow pipeline (Oxford Nanopore).
