## Supplementary Figure S1 for "Hospital wastewater surveillance for SARS-CoV-2 identifies intra-hospital dynamics of viral transmission and evolution"

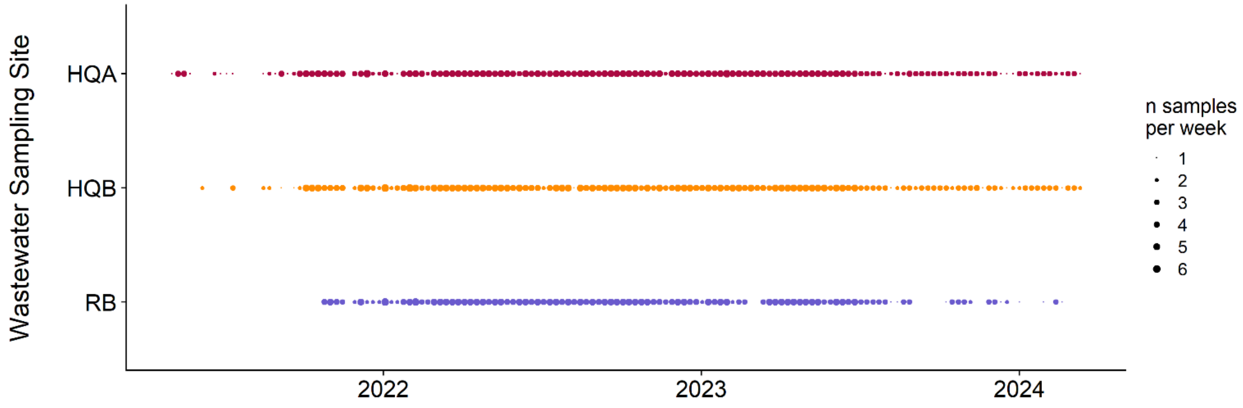


**Supplementary Figure S1. Hospital and research building sampling overview.** For each sampling site (y-axis) over each week of our sampling period (x-axis) from May 2021 through March 2024, points are scaled by the number of samples collected per week. Abbreviations: HQA = hospital quadrant A, HQB = hospital quadrant B, RB = research building
