## Supplementary Table S1 for "Hospital wastewater surveillance for SARS-CoV-2 identifies intra-hospital dynamics of viral transmission and evolution"

**Supplementary Table S1. Weekly wastewater sampling frequency.**

| WASTEWATER SAMPLING SITE | START DATE | END DATE | TOTAL WEEKS OF SAMPLING | NUMBER OF SAMPLES COLLECTED PER WEEK | NUMBER OF WEEKS[1] (% OF TOTAL WEEKS) |
|---|---|---|---|---|---|
| **RB** | 10/26/2021 | 2/21/2024 | 121 | 0 | 14 (12%) |
| | | | | 1 | 7 (6%) |
| | | | | 2 | 10 (8%) |
| | | | | 3 | 22 (18%) |
| | | | | 4 | 19 (16%) |
| | | | | 5 | 47 (39%) |
| | | | | 6 | 2 (2%) |
| **HQB** | 6/9/2021 | 3/13/2024 | 144 | 0 | 10 (7%) |
| | | | | 1 | 7 (5%) |
| | | | | 2 | 15 (10%) |
| | | | | 3 | 35 (24%) |
| | | | | 4 | 23 (16%) |
| | | | | 5 | 52 (36%) |
| | | | | 6 | 2 (1%) |
| **HQA** | 5/5/2021 | 3/13/2024 | 149 | 0 | 7 (5%) |
| | | | | 1 | 14 (9%) |
| | | | | 2 | 15 (10%) |
| | | | | 3 | 26 (17%) |
| | | | | 4 | 32 (22%) |
| | | | | 5 | 53 (36%) |
| | | | | 6 | 2 (1%) |

[1] The number of weeks where *n* samples were collected throughout the sampling period are shown for each sampler.
