## Supplementary Table S2 for "Hospital wastewater surveillance for SARS-CoV-2 identifies intra-hospital dynamics of viral transmission and evolution"

**Supplementary Table S2. Lead-lag correlation analysis between wastewater viral concentration and confirmed clinical cases in HQA with shedding dynamics sensitivity analysis, with and without normalization by crAssphage concentration**

| CROSS-CORRELATION MODEL | Wastewater Data Only[1] $t_0$ (cases) ~ $t_0$ (WW) + $t_{-7}$ (WW) | | | | Normalized Wastewater Data Only[2] $t_0$ (cases) ~ $t_0$ ($WW_{norm}^2$) + $t_{-7}$ ($WW_{norm}$) | | | |
|---|---|---|---|---|---|---|---|---|
| ASSUMED VIRAL SHEDDING WINDOW[3] | **3d** | **7d** | **10d** | **14d** | **3d** | **7d** | **10d** | **14d** |
| HQA model $R^2$ | 0.33 | 0.30 | 0.30 | 0.33 | 0.32 | 0.29 | 0.29 | 0.32 |
| HQA model *p*[4] | *** | *** | *** | *** | *** | *** | *** | *** |
| *p* ($t_0$ WW or $WW_{NORM}$) | ** | * | * | * | * | * | * | * |
| *p* ($t_{-7}$ WW or $WW_{NORM}$) | *** | *** | *** | *** | *** | *** | ** | *** |

[1] Represents a subset of data from Table 1 with available concurrent SARS-CoV-2 and crAssphage viral concentration data (n=60 weeks; January 1, 2023 – March 1, 2024)

[2] WWnorm = wastewater viral concentration normalized by coincident crAssphage concentration using the formula:

Normalized wastewater viral load = wastewater viral load / - log(crassphage viral load)

[3] As calculated using hospital location data from patient electronic medical records; viral shedding window expressed in days after last confirmed clinical positive SARS-CoV-2 test

[4] p<0.05 (*); p<0.01 (**); p<0.001 (***)
