## Supplementary Table S3 for "Hospital wastewater surveillance for SARS-CoV-2 identifies intra-hospital dynamics of viral transmission and evolution"

**Supplementary Table S3. Lead-lag correlation analysis between wastewater viral concentration and confirmed clinical cases with sampling frequency sensitivity analysis**

| CROSS-CORRELATION MODEL | **Wastewater Data Only** $t_0$ (cases)[1] ~ $t_0$ (WW) + $t_{-7}$ (WW) | | |
|---|---|---|---|
| OBSERVATIONS PER WEEK (N) | **1** | **2** | **3** |
| HQA model $R^2$ | 0.01 | 0.01 | 0.13 |
| HQA model *p*[2] | 0.24 | 0.19 | * |
| *p* ($t_0$ WW) | 0.36 | 0.38 | 0.1 |
| *p* ($t_{-7}$ WW) | 0.15 | 0.13 | * |
| HQB model $R^2$ | 0.44 | 0.52 | 0.55 |
| HQB model *p*[2] | *** | *** | *** |
| *p* ($t_0$ WW) | *** | *** | *** |
| *p* ($t_{-7}$ WW) | *** | *** | *** |

[1] As calculated using hospital location data from patient electronic medical records; assumed viral shedding window of 14 days after first confirmed clinical positive SARS-CoV-2 test

[2] $p<0.05$ (*); $p<0.01$ (**); $p<0.001$ (***)
